# Orthogonal validation of PD Biomarkers: Multi-platform proteomics profiling of CSF, Plasma, and Urine confirms DDC as a consistent candidate

**DOI:** 10.1101/2025.09.25.25336658

**Authors:** Ravindra Kumar, Aleksandra Beric, Daniel Western, Zining Yang, Wenjing Lin, Jigyasha Timsina, Carlos Cruchaga, Laura Ibanez

## Abstract

**Background:** High throughput proteomics has enabled hypothesis free biomarker discovery. However, differences in sample sizes, biological fluid, and quantification technologies have limited replication and validation of the results, and studies on the cross-platform variability are lacking. Here, we present the first orthogonal validation across three platforms in Parkinson’s disease (PD) to understand the technical and biological challenges of proteomic studies.

**Methods:** We have leveraged publicly available proteomic data from cerebrospinal fluid (CSF), plasma, and urine within the Parkinson’s Progression Markers Initiative (PPMI) cohort, generated using SomaScan5K (CSF), mass spectrometry (MS; CSF, plasma, and urine), and Olink Explore (CSF and plasma). Across platforms, we compared 375 proteins that were consistently quantified. We performed differential abundance analysis comparing PD versus healthy controls followed by sensitivity analyses (mutation carriers, at-risk participants, longitudinal analyses) to further understand the findings.

**Results:** In CSF, we found significant correlations between effect sizes from the 375 proteins quantified by SomaScan5K and MS (ρ=0.42, p=2.60×10 □ □), as well as SomaScan5K and Olink Explore (ρ=0.15, p=3.15×10□^3^) while MS and Olink Explore showed no significant correlations in CSF or plasma. Orthogonal validation identified two proteins (DLK1, GSTA3) replicated between SomaScan5K and Olink Explore and seven proteins (ALCAM, CHL1, CNDP1, NCAM2, PEBP1, PTPRS, SCG2) replicated between MS and SomaScan5K. No proteins replicated between MS and Olink Explore in CSF or plasma. DDC showed consistent dysregulation across analyses. In CSF (Olink Explore), it was dysregulated in PD participants (beta=0.79, p=8.49×10^−16^), and in at-risk individuals (beta=0.64, p=1.41×10^−7^) including those with hyposmia (beta=0.70, p=2.13×10^−5^) and REM Sleep Behavior Disorder (beta=0.52, p=1.00×10^−3^). In urine, DDC was higher in at-risk individuals (beta=0.43, p=7.28×10^−5^), driven by *LRRK2*^+^ at-risk participants (beta=0.59, p=1.74×10^−6^), as well as in symptomatic mutation carriers, *LRRK2*^+^ (beta=0.68, p=9.08×10^−8^), and *GBA*^*+*^ (beta=0.28, p=0.04).

**Conclusions:** Biologically, these findings add further evidence that DDC has strong potential as a biomarker. Methodologically, our findings emphasize that platform selection can introduce more variance than that originating from disease status, which limits the reproducibility across technologies. This highlights the challenges and importance of cross-platform validation in proteomic biomarker research, and the translation of those discoveries to the clinic.

## Background

Parkinson’s disease (PD) is the second most common neurodegenerative disorder[1] after Alzheimer’s disease, affecting more than six million people worldwide.[2-5] This number has increased 2.5 times over the past 30 years, making PD one of the leading causes of neurological disability.[6] It is clinically characterized by motor and non-motor features such as bradykinesia, rigidity, and tremors. Pathologically, it is defined by the progressive degeneration of dopaminergic neurons in the substantia nigra and accumulation of misfolded alpha-synuclein within Lewy bodies in the brain.[7-9] PD diagnosis currently relies on clinical history and symptoms, as no clinically useful biomarker is available yet.[10, 11] Recently, the CSF alpha-synuclein seed amplification assay (SAA) has emerged as a promising biomarker, but some challenges still remain. Main drawbacks of SAA are its invasiveness, as it is currently most predictive in CSF, and dependence on quantification instead of positive/negative result.[12, 13] Thus, the search for biomarkers, especially early ones, able to detect PD risk continues. High-throughput proteomics technologies such as mass spectrometry (MS), aptamer-based SomaScan, and proximity extension-based Olink have advanced biomarker discovery by enabling high sensitivity and reproducible quantification of proteins across diverse biological samples. Since different platforms utilize different quantification technologies, multi-panel approaches allow for the precise measurement of protein abundance and facilitate the identification of novel potential biomarkers.

Currently, three main quantitative protein measurement technologies (MS, SomaScan, and Olink Explore) are available. MS is based on mass-to-charge ratio of ionized peptides, while SomaScan and Olink Explore are affinity-based technologies. SomaScan uses aptamers (short nucleic acids) to bind and detect target proteins, whereas Olink Explore is an immunoassay-based proximity extension assay using two antibodies.[14-18] In the past few years, numerous studies have utilized MS[19-26], SomaScan[9, 27-30], and Olink Explore[7, 31-36] in PD research, but orthogonal comparison studies have been limited.[37] In 2023, Tsukita *et al*.[37] compared SomaScan with MS in CSF samples. Their study included 4,071 proteins from 420 samples from the Parkinson’s Progression Marker Initiative (PPMI; N_PD_=279, N_Control_=141) measured using SomaScan, and 65 CSF samples (N_PD_=34, N_Control_=31) from the LRRK2 Cohort Consortium (LCC) measured using MS (1,556 proteins). They identified 880 proteins common in both datasets and five orthogonally replicated in both. Rutledge *et al*.[8] in 2024 compared SomaScan and Olink Explore quantifications with MS, across multiple biofluids. They analyzed a total of 4,877 samples from CSF, plasma, and urine from the PPMI and Standford PD cohorts and observed that DOPA decarboxylase (DDC) was the only protein that was consistently upregulated in CSF and urine of treatment-naïve PD and at-risk PD participants, as quantified by all three-proteomic platforms. A more recent and smaller study measuring DDC using Olink Explore suggests that changes in DDC are due to treatment and not pathology.[34]

Overall, and despite progress in the field, research published in recent years has not comprehensively compared high-throughput proteomic quantifications in the same cohort. Most studies focus on a single biofluid or platform; and even if they include multi-platform analyses, very few studies directly compare affinity-based technologies Olink Explore and SomaScan with MS-based platform. In fact, only one study to date has incorporated all three technologies[8], but they did not perform an orthogonal validation. Given the complex and heterogenous nature of PD, understanding the role of quantification technology is crucial in identifying reliable biomarkers and translating them to the clinic.

To address the aforementioned limitations, we conducted an orthogonal analysis integrating publicly available proteomic data from three biofluids across three proteomic technologies within the PPMI cohort using the same statistical model. This study represents the first effort to analyze and compare multi-fluid, cross-platform proteomic data and attempt orthogonal validation of putative PD biomarkers.

## Methods

### Study design

We leveraged publicly available proteomic quantifications from CSF, plasma, and urine, generated within the PPMI cohort, using three high-throughput proteomic platforms: SomaScan5K, MS, and Olink Explore. After quality control (QC), 3,380 CSF, 1,426 plasma, and 817 urine samples were included in downstream analyses. All urine samples were quantified using MS. CSF was quantified using SomaScan5K (N=1,020), MS (N=1,811), and Olink Explore (N=549), while plasma was quantified using MS (N=792) and Olink Explore (N=634; Figure 1). Further, we identified the proteins that were quantified by all three technologies and performed orthogonal validation using the same minimally adjusted statistical model. We compared healthy control (HC) and PD participants (regardless of mutation status) to identify differentially accumulated proteins through cross-sectional analyses using baseline visit. To further contextualize our findings, we performed several sensitivity analyses: (i) cross-sectional analyses using participants’ last visit, (ii) longitudinal analyses using those participants who had at least two time points, (iii) cross-sectional analyses of PD participants with *GBA*^*+*^ and *LRRK2*^*+*^ mutations using baseline visits, and (iv) cross-sectional analyses with at-risk participants, including at-risk mutation carriers, as well as participants with hyposmia or Rapid Eye Movement (REM) Sleep Behavior Disorder (RBD), using baseline visits. To biologically understand our findings, we performed pathway analyses. The study was approved by the Washington University in Saint Louis Institutional Review Board (IRB ID 201701124 and 202004010).

**Figure 1.**
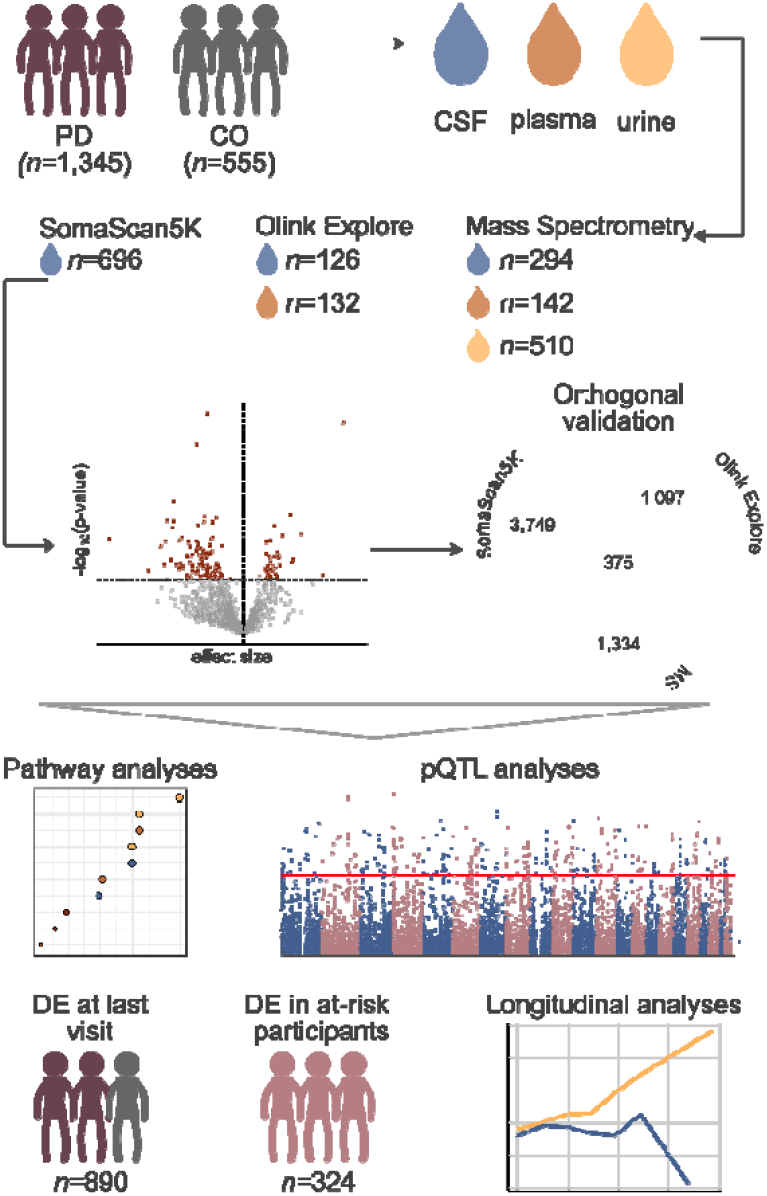
Infographic of the study design depicting collection of CSF, plasma and urine from healthy control and PD participants, protein quantification via one of the three high-throughput proteomic platforms (SomaScan5K, Olink Explore, and MS), summary of main analyses including differential accumulation and orthogonal validation, followed by biological contextualization of the results via pathway, as well as sensitivity analyses.

### Sample description

All the data included in the present work are publicly available from PPMI and can be accessed via AMP-PD **(https://amp-pd.org/)** or LONI by qualified investigators. PPMI is a global initiative that has enrolled over 4,000 participants to date, including HC, individuals diagnosed with PD that can be carriers of a mutation known to be causally associated with PD[38, 39], and participants at-risk of developing PD who show no symptoms, but carry a PD-related mutation, or suffer from RBD, or hyposmia, both syndromes associated with increased PD risk.[40, 41]

We included 2,835 samples corresponding to 1,214 unique Non-Hispanic White (NHW) participants from PPMI. Across different fluids and platforms sample size was variable, but the demographic and clinical profiles remained consistent (Table 1).

**Table 1.** Demographic and clinical characteristics of baseline participants.

| Platform | Fluid | Disease status | Unique individuals | Mean age (*SD) | Female (N, %) | Mean UPDRS-III (*SD) | Mean MoCA (*SD) |
| --- | --- | --- | --- | --- | --- | --- | --- |
| SomaScan5K | CSF | HC | 155 | 61.26<br>($\pm 11.05$) | 57<br>(36.77%) | 1.21 ($\pm 2.21$) | 28.20<br>($\pm 1.13$) |
| | | PD | 541 | 62.21<br>($\pm 9.36$) | 213<br>(39.37%) | 21.40 ($\pm 9.70$) | 26.90<br>($\pm 2.48$) |
| | | At-Risk | 324 | 61.51<br>($\pm 7.35$) | 192<br>(59.26%) | 2.68 ($\pm 3.81$) | 26.90<br>($\pm 2.24$) |
| MS | CSF | HC | 88 | 62.52<br>($\pm 10.18$) | 29<br>(32.95%) | 1.27 ($\pm 2.21$) | 28.20<br>($\pm 1.11$) |
| | | PD | 206 | 61.45<br>($\pm 9.53$) | 71<br>(34.47%) | 20.40 ($\pm 9.04$) | 27.30<br>($\pm 2.11$) |
| | | At-Risk | 60 | 63.08<br>($\pm 6.82$) | 31<br>(51.67%) | 2.92 ($\pm 4.10$) | 26.60<br>($\pm 2.42$) |
| | Plasma | HC | 48 | 61.23<br>($\pm 10.94$) | 14<br>(29.17%) | 1.17 ($\pm 1.92$) | 28.20<br>($\pm 1.06$) |
| | | PD | 94 | 61.24<br>($\pm 9.98$) | 31<br>(32.98%) | 19.70 ($\pm 8.69$) | 27.20<br>($\pm 2.07$) |
| | | At-Risk | 20 | 63.09<br>($\pm 4.99$) | 10<br>(50.00%) | 2.80 ($\pm 3.86$) | 26.80<br>($\pm 2.48$) |
| | Urine | HC | 106 | 61.75<br>($\pm 10.56$) | 45<br>(42.45%) | 1.36 ($\pm 2.53$) | 28.20<br>($\pm 1.13$) |
| | | PD | 404 | 62.68<br>($\pm 9.35$) | 180<br>(44.55%) | 21.90 ($\pm 9.99$) | 26.80<br>($\pm 2.48$) |
| | | At-Risk | 307 | 62.09<br>($\pm 7.23$) | 173<br>(56.35%) | 2.60 ($\pm 3.65$) | 26.80<br>($\pm 2.29$) |
| Olink Explore | CSF | HC | 79 | 61.02<br>($\pm 11.12$) | 26<br>(32.91%) | 1.13 ($\pm 2.11$) | 28.20<br>($\pm 1.13$) |
| | | PD | 47 | 61.76<br>($\pm 9.86$) | 12<br>(25.53%) | 20.90 ($\pm 9.68$) | 27.30<br>($\pm 1.84$) |
| | | At-Risk | 36 | 68.64<br>($\pm 6.50$) | 13<br>(36.11%) | 4.22 ($\pm 4.45$) | 26.00<br>($\pm 3.50$) |
| | Plasma | HC | 79 | 61.49<br>($\pm 10.53$) | 25<br>(31.65%) | 1.06 ($\pm 1.91$) | 28.20<br>($\pm 1.15$) |
| | | PD | 53 | 61.92<br>($\pm 11.37$) | 15<br>(28.30%) | 21.80 ($\pm 9.59$) | 27.10<br>($\pm 2.17$) |
| | | At-Risk | 42 | 67.87<br>($\pm 6.32$) | 13<br>(30.95%) | 3.93 ($\pm 4.28$) | 26.30<br>($\pm 2.67$) |

\*Standard deviation

SomaScan5K was used to generate proteomic data on 4,775 aptamers in 1,020 CSF samples, all from baseline visits. MS was used to obtain measurements for 291, 185, and 1,819 peptides in 1,811 CSF, 792 plasma, and 817 urine samples respectively corresponding to 990 unique individuals with longitudinal assessments available for CSF and plasma only. Olink Explore was leveraged to quantify 1,459 proteins in 549 CSF samples and 1,450 proteins in 634 plasma samples, corresponding to 253 unique individuals with longitudinal assessments (Table 2). All the longitudinal data were collected every six months for both CSF and plasma. While each platform uses samples from PPMI participants, there is limited overlap between participants and samples across fluids and technologies (Supplementary Figure 1).

**Table 2.**
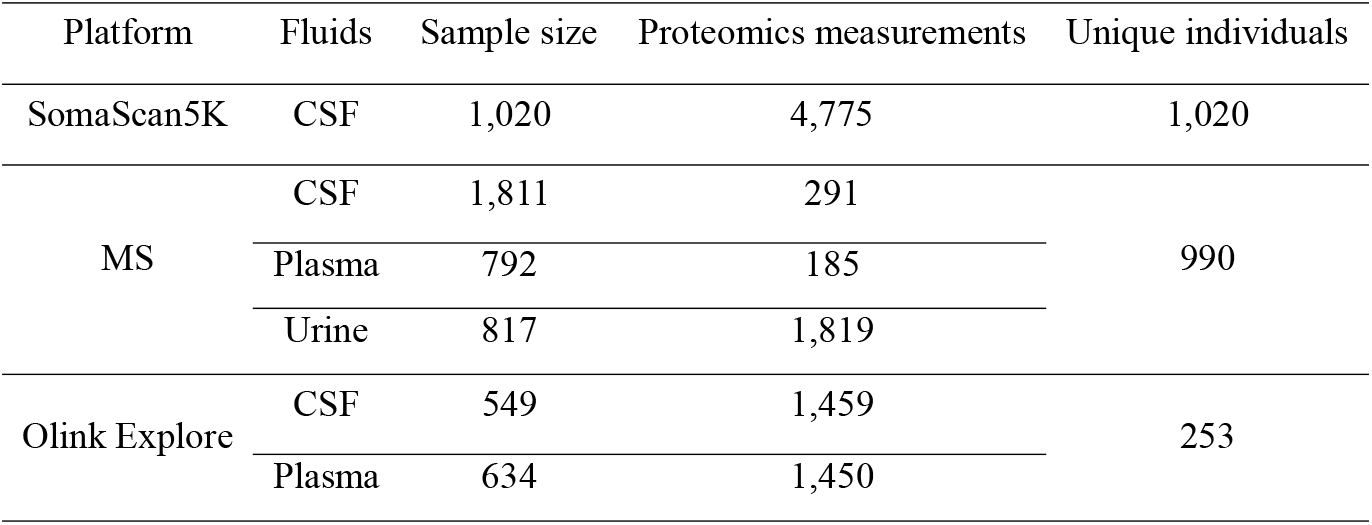
Sample information for different fluids and platforms.

### Data processing and QC

Data from all three platforms (SomaScan5K, MS and Olink Explore) underwent similar QC steps. First, outlier measurements were identified using interquartile range (IQR) and treated as missing if expression values were below *Q1-1*.*5IQR* or above *Q3+1*.*5IQR*. Then, the call rate, the percent of non-missing values, was calculated for each aptamer/peptide/protein and each sample. Any aptamers/peptides/proteins with call rate below 65% were removed. Call rates were then recalculated for the remaining features, and samples with a call rate below 65% were subsequently excluded. Further, we applied a more stringent threshold, removing any aptamers/peptide/proteins with call rate below 85%, before recomputing call rate by sample and excluding any sample with call rate below 85%. All values obtained with SomaScan5K or MS were scaled using z-scores (Supplementary Figure 2). Olink Explore data was not scaled, since the measurements are reported as Normalized Protein eXpression (NPX), a normalized unit on a log_2_ scale. Next, principal component analyses (PCA) were used to identify and remove sample outliers. Outliers were defined as being more than three standard deviations away from the means of either of the two first PC (Supplementary Figure 3).

After QC, we retained 4,775 aptamers in the SomaScan5K CSF data, corresponding to 4,093 unique protein measurements, from 1,020 samples (N_HC_=155, N_PD_=541, and N_At-risk_=324). In MS, we retained 291 peptides from 1,811 CSF samples, corresponding to 447 unique participants (N_HC_=459, N_PD_=1,064, and N_at-risk_=288) with a maximum of 11 visits. In MS plasma, we kept 185 peptides from 792 samples corresponding to 167 unique participants (N_HC_=209, N_PD_=479, and N_at-risk_=104) with a maximum of ten visits, while in urine, we retained 1,819 peptides from 817 baseline samples (N_HC_=106, N_PD_=404, and N_at-risk_=307). Finally, in Olink Explore, 1,459 proteins from 549 CSF samples corresponding to 233 unique participants (N_HC_=297, N_PD_=125, and N_at-risk_=127) along nine visits were retained after QC, whereas in plasma, 1,450 proteins from 634 samples (N_HC_=299, N_PD_=148, and N_at-risk_=187) from 250 unique participants spanning up to ten visits remained.

### Statistical analyses

#### Cross-sectional analysis

We performed cross-sectional differential abundance analysis using linear regression with the *lm()* function from the base R package. To identify proteins that are differentially abundant in PD cases (regardless of mutation status) compared to HC, we used baseline measurements, available across all platforms and tissues. We used a minimally adjusted model adding age at draw and sex as covariates for all fluids and quantification technologies. All p-values were corrected for multiple testing using the Benjamini-Hochberg correction (FDR). Proteins were considered differentially accumulated if the FDR corrected p-value was lower than 0.05. Pathway enrichment analysis was performed using the ClusterProfiler R package.[42] In CSF and plasma from MS and Olink Explore, enrichment was based on nominal significant proteins due to limited peptide/protein detection. In contrast, CSF from SomaScan5K and urine from MS were performed using FDR-based differentially accumulated proteins.

#### Sensitivity analyses

To better understand the findings, we performed differential abundance analyses using the last-visit available for each participant, in an attempt to maximize the biological differences between PD participants and HC by selecting participants with longer disease duration. Next, and because we performed our main analyses with all PD cases regardless of mutation status, we assessed the impact of genetic background on protein accumulation at baseline and investigated whether our results were driven by idiopathic PD or PD participants who were *LRRK2*^+^ and *GBA*^+^ mutation carriers by comparing each group to HC separately. Additionally, we leveraged baseline data from at-risk participants (those who do not have a PD diagnosis but suffer from RBD, hyposmia, or carry mutations in *LRRK2*^+^ or *GBA*^+^ genes), to test whether any of the identified proteins were differentially accumulated before symptom onset. Due to insufficient mutation data for samples quantified via Olink Explore, sensitivity analyses using measurements from this platform were not performed. Sensitivity analyses with *SNCA*^*+*^ mutation carriers were not conducted due to the low number of participants with proteomic information that carry mutations in the *SNCA* gene (N=9).

Additionally, due to the longitudinal nature of some of the data, we leveraged the longitudinal characteristics of the PPMI study and applied linear mixed models to assess whether the proteins identified in the previous section change significantly over time. We included participants with at least two clinical visits and the corresponding proteomic measurements available after QC. All analyses were adjusted by protein levels at baseline, sex, and age at baseline; *protein×time* was added as the interaction term and participant ID as the random effect term. We used *lmer* function from lme4 R package[43] for differential analysis and model generation. For all available tissues and platforms, pathway enrichment analysis was performed using ClusterProfiler R package[42] utilizing nominally significant findings.

#### pQTL mapping

We performed protein quantitative trait loci (pQTL) mapping for baseline CSF measurements combining genomic data and proteomic data derived from each of the three protein measurement technologies. We removed any cryptic and non-cryptic related samples using identity-by-descent filtering (using a PI_HAT threshold of 0.25 to define related pairs) and selected only samples form NHW based on principal component analysis, anchored using the 1000 Genomes Project samples of known ancestry. After all genotype filtering criteria, 117, 742, and 288 participants were included in the pQTL analysis for Olink Explore, SomaScan5K, and MS, respectively.

The pQTL mapping was performed using a linear regression approach in PLINK2[44] using age, sex, and the first ten genetic principal components as covariates. To account for the varying sample sizes between the protein platforms, variant missingness and allele counts were calculated independently for each platform; all variants with missingness ≥ 0.1 or minor allele count (MAC) < 10 were excluded from the analyses. In total, 7,660,921 variants passed QC for the MS analyses, 6,265,584 for the Olink analyses, and 9,258,707 for the SomaScan analyses. Significant pQTLs were identified using the genome-wide significance threshold (p<5×10^−8^) and are denoted using the most-significant variant within each pQTL locus.

## Results

### Quantification technology has a large effect on the findings

To orthogonally validate proteomic findings using multiple high-throughput quantification platforms and different fluids, we first identified 375 proteins shared by all three platforms regardless of tissue (Figure 2A). Given the limited overlap of samples measured with the three technologies (Supplementary Figure 1A), we performed differential abundance analyses at baseline visit, followed by pair-wise comparisons of effect sizes for the 375 shared proteins using Spearman correlation. In CSF, we observed significant correlation between the 71 proteins quantified by SomaScan5K and MS (ρ=0.42, p=2.60×10□ □), as well as SomaScan5K and Olink Explore (ρ=0.15, p=3.15×10□^3^; 373 proteins). However, correlations between MS and Olink Explore were not significant in either CSF (71 proteins) or plasma (20 proteins) (Figure 2B, Supplementary Figure 4, and Supplementary Table 1).

**Figure 2.**
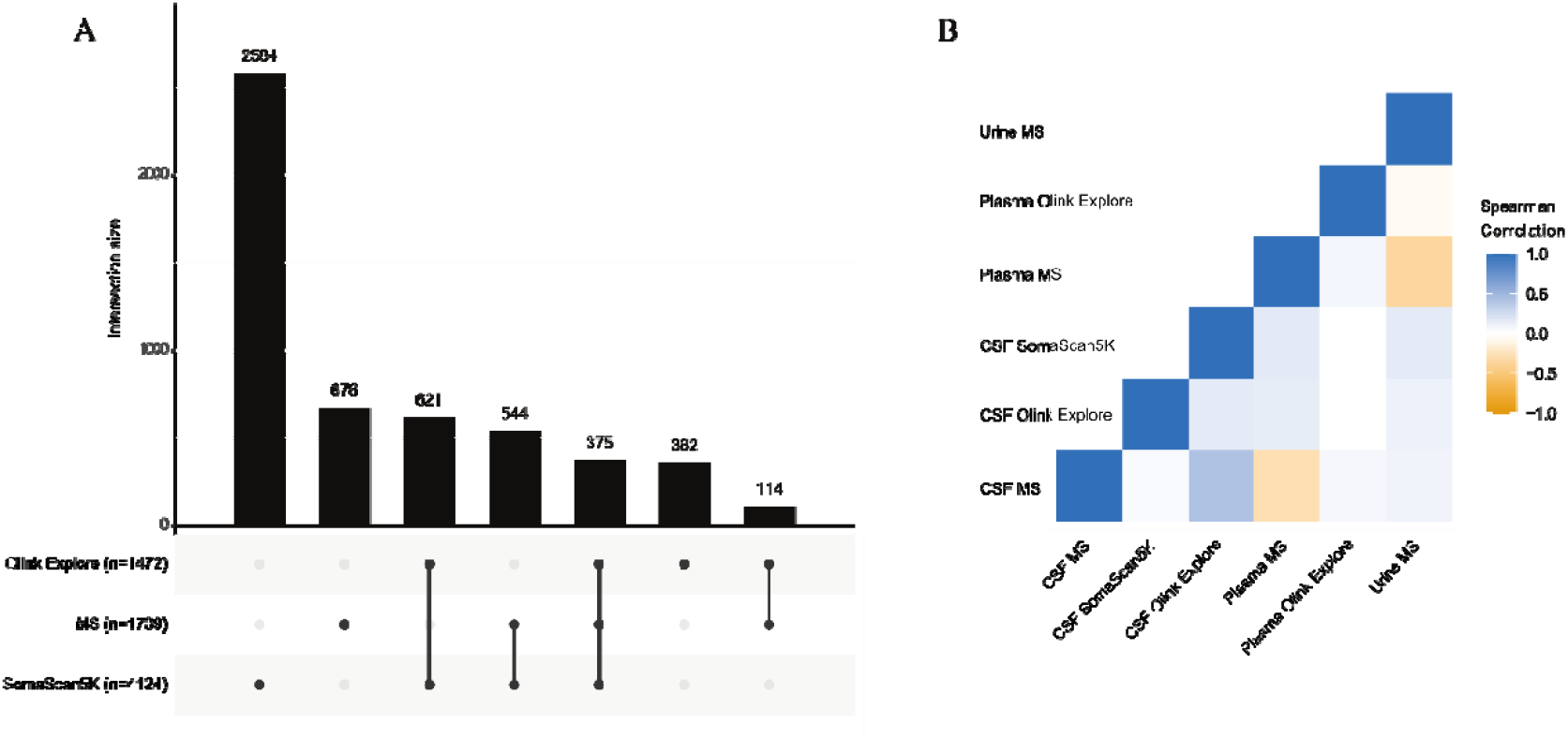
Proteins quantified by the different technologies, regardless of fluid, and correlation of effect sizes obtained from differential analyses using a minimally adjusted statistical model. **A.** Upset plot illustrating proteins quantified by the different technologies, regardless of tissue, and their overlap and **B**. heatmap depicting the correlation of effect sizes for commonly quantified proteins across various fluids and technologies. *Represents significant correlation.

We then attempted orthogonal validation of proteins. Since not all analyses resulted in proteins that were significant after FDR correction, we performed the orthogonal validation with nominally significant findings and identified five proteins that showed nominal significance in CSF at baseline visit for at least two technologies: CPVL (SomaScan5K: beta=-0.30, p=6.77×10^−4^; Olink Explore: beta=0.16, p=0.03), DLK1 (SomaScan5K: beta=-0.44, p=9.13×10^−7^; Olink Explore: beta=-0.13, p=0.04), DNER (SomaScan5K: beta=-0.18, p=0.09; Olink Explore: beta=0.03, p=0.04), GP6 (SomaScan5K: beta=0.25, p=5.00×10^−3^; Olink Explore: beta=-0.13, p=4.96×10^−2^), and GSTA3 (SomaScan5K: beta=-0.26, p=5.73×10^−3^; Olink Explore: beta=-0.19, p=0.04; Supplementary Table 2). Among these, DLK1, and GSTA3 were reduced in PD cases compared to controls in both platforms, while CPVL, DNER and GP6 showed opposite direction of effect, preventing the orthogonal validation. Similarly, we found seven proteins, ALCAM (SomaScan5K: beta=-0.34, p=6.52×10^−5^; MS: beta=-0.25, p=0.04), CHL1 (SomaScan5K: beta=-0.43, p=6.92×10^−7^; MS: beta=-0.34, p=5.32×10^−3^), CNDP1 (SomaScan5K: beta=-0.27, p=2.36×10^−3^; MS: beta=-0.25, p=0.04), NCAM2 (SomaScan5K: beta=-0.28, p=1.46×10^−3^; MS: beta=-0.26, p=0.04), PEBP1 (SomaScan5K: beta=-0.28, p=1.32×10^−3^; MS: beta=-0.26, p=0.04), PTPRS (SomaScan5K: beta=-0.30, p=3.20×10^−4^; MS: beta=-0.34, p=9.25×10^−3^), and SCG2 (SomaScan5K: beta=-, p=1.90×10^−6^; MS: beta=-0.39, p=9.25×10^−4^) nominally dysregulated at baseline in CSF samples measured with MS and SomaScan5K in the same direction of effect (Supplementary Table 3). No proteins were found to be orthogonally replicated between MS and Olink Explore in either CSF or plasma.

### DDC and other urine specific proteins are altered in PD cases

DDC was differentially accumulated in both CSF from Olink Explore and urine from MS. It was identified as the most differentially upregulated protein in PD participants compared to HC in CSF (beta=0.66, p=6.54×10^−10^) from Olink Explore (Supplementary Table 4) as reported previously.[8, 31, 35, 45] DDC was also found to be differentially upregulated in urine (beta=0.37, p=4.82×10^−4^, Supplementary Table 5), consistent with earlier findings[8]. In CSF quantified with Olink Explore, the mean expression of DDC was higher in PD (p=2.22×10^−16^), while in urine from MS, mean expression was higher in at-risk participants (p=3.00×10^−7^) than in PD (Supplementary Figure 5). To further elucidate the role of DDC, we conducted several sensitivity analyses by comparing HC with PD participants at their last visit, HC with at-risk participants and at-risk with PD participants with baseline measurements. We observed that DDC was significantly upregulated in the last visit of PD cases (beta=0.79, p=8.49×10^−16^) compared to HC and in at-risk participants (beta=0.64, p=1.41×10^−7^) compared to HC at their baseline (Supplementary Table 4). Further, we found an increase in DDC levels in at-risk participants with hyposmia (beta=0.70, p=2.13×10^−5^) and RBD symptoms (beta=0.54, p=1.00×10^−3^, Supplementary Table 4) in their baseline measurements. We also observed that DDC levels increased over time in CSF in PD participants relative to HC (Supplementary Figure 6). In sensitivity analyses of urine, we observed that DDC was differentially upregulated in at-risk participants (beta=0.43, p=7.28×10^−5^) compared to HC at baseline. When performing analyses on mutation carriers, increased DDC levels were observed in PD participants with *GBA* mutations (beta=0.28, p=0.04) and *LRRK2* mutations (beta=0.68, p=9.08×10^−8^) compared to HC, and at-risk participants with *LRRK2* mutation (beta=0.59, p=1.74×10^−6^) (Supplementary Table 5).

To further investigate whether DDC was associated with motor symptoms, we assessed the relationship between DDC from Olink Explore and MS and UPDRS III. We found a significant correlation (ρ=0.33, p=6.37×10^−4^) in CSF and a weak non-significant correlation (ρ=0.08, p=0.10) in urine (Figure 3A and B). Given the quantification of DDC in the Olink Explore panel, we checked correlation with the samples that were quantified with both technologies (N=22). No significant correlation was observed (ρ=0.28, p=0.20) between DDC quantifications, most likely driven by the small number of overlapping samples (Figure 3C).

**Figure 3.**
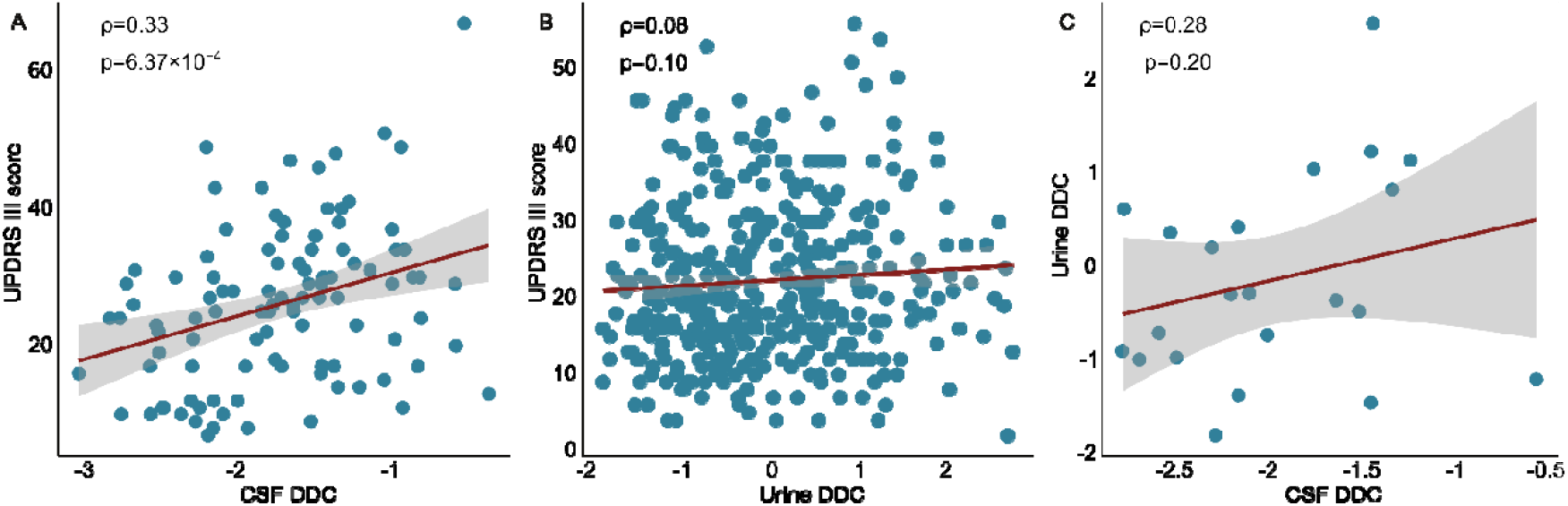
Correlation of normalized DDC expression levels in PD participants. (A) correlation between UPDRS III score and DDC expression in CSF; (B) correlation between UPDRS III score and DDC expression in urine; (C) correlation of DDC expression in CSF measured by Olink Explore and in urine measured by MS.

### There are no shared CSF proteomic signatures across all quantification technologies

We identified differentially accumulated proteins within each individual technology at baseline visit and performed pairwise comparisons. In CSF from SomaScan5K, we identified 834 aptamers corresponding to 775 proteins, with the top ones being LPO (beta=-0.71, p=4.30×10^−18^), HAMP (beta=0.58, p=2.84×10^−11^), WIFI (beta=-0.50, p=2.98×10^−9^), and NETO1 (beta=-0.52, p=3.53×10^−9^, Supplementary Table 6, Supplementary Figure 7A). When performing sensitivity analyses, out of the 834 aptamers, 800 were dysregulated in sporadic PD, 370 in PD cases that carry a *GBA*^*+*^ mutation, 628 in PD cases with *LRRK2*^*+*^ mutation, and 191 in at-risk participants. In participants at risk, 306 aptamers were dysregulated in at-risk participants carrying a *GBA*^*+*^ mutation, 128 in at-risk participants carrying *LRRK2*^*+*^ mutation, and 94 were common in both non-symptomatic mutation carrier groups (Supplementary Table 6).

In CSF quantified with MS, no significant peptides were identified, but 47 peptides were nominally significant, including VGF (beta=-0.46, p=3.26×10^−4^), CDH13 (beta=-0.44, p=3.76×10^−4^), SCG2 (beta=-0.39, p=9.26×10^−4^), APOA1 (beta=0.42, p=1.17×10^−3^) (Supplementary Table 7, Supplementary Figure 7B). Of those, 29 remained nominally significant at last visit, three in *GBA*^*+*^ mutation carriers, five in *LRRK2*^*+*^ mutation carriers, and four in at-risk participants compared to HC. When evaluating the different at-risk groups, four peptides were nominally significant in at-risk participants carrying *LRRK2* mutations, five in at-risk participants with hyposmia, and seven showed differences in the rate of change. In CSF quantified using Olink Explore, only one protein, DDC (beta=0.67, p=6.54×10^−10^), was differentially accumulated as previously described[8], while 50 were nominally significant (Supplementary Table 4, Supplementary Figure 7C).

To biologically contextualize our findings, we conducted pathway enrichment analyses for findings from each proteomic platform separately. The analysis leveraging the 834 differentially accumulated aptamers in SomaScan5K identified 16 significantly enriched KEGG pathways including cell adhesion molecule (p=1.40×10^−13^), axon guidance (p=4.32××10^−13^), cytokine-cytokine receptor interaction (p=8.31×10^−13^), MAPK signaling (p=5.75××10^−7^), and JAK-STAT signaling pathway (p=3.82×10^−4^; Supplementary Table 8). Since few proteins differentially accumulated in CSF from MS and Olink Explore, we instead used all proteins with a nominal p-value for pathway enrichment analysis. In CSF from MS, 47 nominally significant proteins mapped to four KEGG pathways (Supplementary Table 9) while in CSF from Olink Explore, 50 proteins mapped 23 pathways (Supplementary Table 10). Of these, one pathway, cell adhesion molecules (p-value: SomaScan5K=1.39×10^−13^, MS=6.18×10^−6^), was enriched in both SomaScan5K and MS, and three pathways, MAPK signaling (p-value: SomaScan5K=5.74×10^−7^, Olink Explore=1.19×10^−4^), JAK-STAT signaling pathway (p-value: SomaScan5K=3.82×10^−4^, Olink Explore=3.79×10^−3^), and viral protein interaction with cytokine and cytokine receptor (p-value: SomaScan5K=1.77×10^−3^, Olink Explore=6.59×10^−3^), were identified in both SomaScan5K and Olink Explore. No pathway was shared across all three platforms.

### Limited shared plasma proteomic signatures between MS and Olink Explore

We also evaluated plasma measurements using MS and Olink Explore. We did not find any differentially accumulated peptides/proteins in MS or Olink Explore. However, nine peptides including APOC4 (beta=-0.48, p=9.84×10^−3^), APOC3 (beta=-0.43, p=0.02), APOC2 (beta=-0.38, p=0.02), and CLEC3B (beta=-, p=0.02) were nominally significant in MS (Supplementary Table 11, Supplementary Figure 7D). Out of those nine peptides, only APOC3 (beta=-0.34, p=0.04) showed significance at last visit analyses, suggesting that the other eight are early changes. Two, FKBP5 (beta=0.80, p=0.02), and VASN (beta=-0.72, p=0.02) were dysregulated in *LRRK2*^*+*^ mutation carriers, suggesting that the results were driven by this subgroup, and two peptides, ATP5PO (beta=-0.10, p=3.54×10^−3^) and FKBP5 (beta=-0.08, p=0.01) showed significant rate of change in the longitudinal sensitivity analyses. In plasma from Olink Explore, 127 proteins, the top three of which are ITGAV (beta=-0.15, p=1.38×10^−4^), HPGDS (beta=-0.22, p=9.88×10^−4^), PRDX6 (beta=-0.38 p=1.02×10^−3^) were nominally significant (Supplementary Table 12, Supplementary Figure 7E). Out of these 127 proteins 28 were dysregulated in last visit analyses and eight in at-risk participants compared to HC. When analyzing at-risk subgroups, seven proteins were already dysregulated in at-risk individuals with hyposmia and six in at-risk with RBD. TFF3 was dysregulated in at-risk participants with both hyposmia (beta=0.20, p=0.02) and RBD (beta=0.26, p=4.82×10^−3^). Finally, out of 127, the rate of change of ten proteins were dysregulated in the longitudinal analyses.

We then used nominally significant proteins to perform pathway enrichment analyses. In plasma from MS, we identified two significantly enriched pathways, cholesterol metabolism (p=3.45×10^−4^) and estrogen signaling pathway (p=2.54×10^−3^; Supplementary Table 13), while from Olink Explore, we identified 13 significantly enriched pathways such as cell adhesion molecules (p=6.45×10^−7^), cytokine-cytokine receptor interaction (p=1.01×10^−3^), and ErbB signaling pathway (p=1.96×10^−3^), (Supplementary Table 14). We did not identify any common pathways in plasma from MS and Olink Explore.

### MS captures unique PD proteomic signatures in urine

Finally, we assessed protein accumulation in urine. Out of 1,819 peptides, 222 peptides, corresponding to 212 unique proteins, were differentially accumulated in PD cases compared to HC (Supplementary Figure 7F, Supplementary Table 5). DPP7 (beta=0.53, p=1.61×10^−7^) and FUCA1 (beta=0.51, p=4.58×10^−7^) were the top upregulated peptides while TNXB_004 (beta=-0.50, p=2.50×10^−6^) and HMCN1_002 (beta=-0.51, p=3.73×10^−6^) were the top downregulated peptides in the urine. In mutation analyses, 222 peptides were dysregulated in PD with *LRRK2*^*+*^ mutation, 53 in PD with *GBA*^*+*^ mutations. Among at-risk participants, 196 peptides were dysregulated in at-risk participants compared to HC, 413 were dysregulated in those with *LRRK2*^*+*^ mutation, 191 in those with *GBA*^*+*^ mutation, and 188 in at-risk participants carrying both *LRRK2*^*+*^ and *GBA*^*+*^ mutations.

Further, we leverage 222 differentially expressed peptides to perform pathway analyses and identified 17 significantly enriched KEGG pathways including lysosome (p=1.50×10^−17^), focal interaction (p=2.20×10^−7^), glycosaminoglycan degradation (p=4.30×10^−7^), renin-angiotensin system (p=5.10×10^−4^; Supplementary Table 15).

### CSF pQTL mapping: Minimal overlap suggests non-equivalent measurements

Given the substantial overlap between the proteomic and genomic data in the PPMI cohort and the relevance of genetics in uncovering potentially causal mechanisms of disease etiology, we also performed protein quantitative trait locus (pQTL) mapping in CSF. After filtering to unique NHW participants, 288 MS, 117 Olink Explore, and 742 SomaScan5K samples were used for pQTL mapping, with each platform analyzed separately. Using a stringent genome-wide significance threshold, we identified 57 pQTLs for proteins measured by MS, 148 for Olink Explore, and 2,346 for SomaScan5k (Supplementary Table 16). These results largely mirror those from the differential abundance analysis, in that both sample size and number of measured proteins likely contribute to differences in pQTL detection. While few proteins are measured in multiple panels, we nonetheless sought to determine the pQTL concordance across protein measurement approaches. Only 16 index pQTL variants were for proteins shared between MS and Olink Explore; they had a non-significant (p=0.18) Pearson correlation of 0.36. Because of the much larger number of proteins in the SomaScan5K platform, the variant overlap was much higher. When comparing pQTL effect sizes between MS and SomaScan5K, a weak but significant (*R*=0.21, p=8.00×10^−3^) correlation was observed. The correlation was much higher between Olink Explore and SomaScan5K (*R*=0.48, p<2.2×10^−16^).

## Discussion

To our knowledge this is the first and largest study to date attempting orthogonal validation of biomarkers using high-throughput technologies in the context of PD. We have analyzed publicly available proteomic data from the PPMI study that includes more than 1,300 individuals and 3,700 samples from three fluids assayed with three proteomic platforms. Due to striking differences in the peptides and proteins measured by each technology, we were unable to orthogonally validate any proteins across three platforms. However, we identified five CSF proteins, CPVL, DLK1, DNER, GP6, and GSTA3, that were significant in SomaScan5K and Olink Explore quantifications, and seven CSF proteins (ALCAM, CHL1, CNDP1, NCAM2, PEBP1, PTPRS, and SCG2) that were significant in the quantifications by SomaScan5K and MS. Notably, among the five CSF proteins identified by SomaScan5K and Olink Explore, three of them, DLK1, DNER, and GP6, are membrane associated proteins suggesting that affinity-based platforms may provide reliable quantification for membrane associated proteins. Moreover, we also observed a modest correlation between SomaScan5K and Olink Explore, consistent with previous reports in plasma[46], and between SomaScan5K and MS in CSF, in contrast to what we observed in QTL analyses, where the overlap was very limited. Overall, our results suggest that the choice of quantification platform is a major source of variability, potentially more than pathological status. This finding does not diminish the strengths of high-throughput technologies for discovery of new proteins involved in disease pathology and can be leveraged either as biomarker and/or therapeutic targets. However, it suggests caution in their interpretation and calls for formal validation prior to translation to the clinic. Thus, large sample sizes, reliance on one platform for quantification of all samples, and further validation in clinical setting with highly accurate measurement technologies are still needed.

We have used the same statistical model with different technologies applied on the PPMI cohort to assess the variability that technological platform introduces. From our results, we derive that the variability could be attributed to several factors that should be considered in the analytical design, including the fact that these platforms utilize distinct detection technologies, with SomaScan5K employing aptamer-based affinity binding, Olink Explore a proximity extension assay (PEA), and MS utilizing a label-free, mass to charge ratio detection.[^47, 48^] These differences in detection mechanisms can influence protein quantification, leading to variations in measured effect size. Secondly, each platform may recognize different epitopes of the same protein, capturing distinct isoforms or post-translational modifications, which could contribute to the observed discrepancy. Further, variability in data normalization and processing techniques across platforms may further impact results, and finally, despite leveraging the same cohort, the sample and participant overlap is limited, which can also affect the success of the orthogonal validation.

Regarding specific proteins, we observed that DDC was significantly upregulated in PD participants compared to HC in CSF using Olink Explore as previously described,[8, 31, 35] with additional elevation observed in urine as in previous analysis[8]. The consistent upregulation of DDC across these fluids suggests a systemic alteration in its expression, potentially reflecting neurodegenerative processes. We observed that DDC was upregulated in PD cases in both baseline and last visit, as well as in at-risk participants compared to HC, particularly in CSF, with stepwise increase from HC to at-risk participants and from at-risk to PD cases. This pattern suggests that DDC may serve as an early indicator of PD disease onset instead of driven by treatment as suggested for Lewy Body Dementia (DLB). [8] Moreover, DDC levels were also significantly increased in participants with hyposmia and RBD, further supporting its potential involvement in early PD pathophysiology. Similar to CSF, DDC level in urine were also elevated in at-risk participants, which suggests that the protein can be used as a non-invasive early indicator of disease onset. Additionally, we observed a significant correlation between DDC level and UPDRS-III in CSF suggesting that higher DDC levels may correspond to greater motor impairment and be useful to follow-up disease progression, provided future studies evaluate the effect of ON/OFF medication status. This also would align with the previous findings linking DDC to DLB. Further studies might elucidate if the elevation is due to disease progression or medication metabolism. In both cases, DDC can be used to track disease evolution and/or therapy engagement. In contrast, DDC levels in urine were not correlated to UPDRS, indicating that urinary DDC may not reliably reflect motor symptom severity despite being useful for early detection. We also identified other promising biomarkers in each biofluids. In CSF, LPO was one of the top hits, for which we also identified a cis pQTL.

This research is subject to several limitations: (i) the exclusive focus on NHW individuals restricts the generalizability of the findings, (ii) the use of publicly available data prevented the comparison of the current capabilities of the different platforms, (iii) variations in sample sizes may impact the statistical power of individual comparisons and affect cross-platform consistency, (iv) difference in sensitivity, detection limits, and dynamic range among technologies can lead to substantial variability in protein detection and quantification, and (v) platform-specific detection chemistries introduce unique biases, particularly affecting the accurate quantification of proteins that are membrane bound, highly glycosylated proteins, or expressed at low abundance.

In summary, this study represents the most comprehensive orthogonal validation effort in PD to date, integrating data across three distinct proteomic platforms and multiple biofluids. Our findings reveal that the selection of proteomic technology substantially influences data variance and biomarker detection, underscoring a fundamental limitation in cross-platform reproducibility. These insights highlight the urgent need for harmonized benchmarking strategies and rigorous validation. Regardless, we have been able to provide additional supporting evidence to the use of DDC for early detection and disease monitoring. By systematically dissecting platform-specific biases while identifying reproducible protein signatures, this work establishes a foundational framework to improve the robustness and generalizability of proteomic biomarkers in neurodegenerative disease.

## Data Availability

PPMI data is available from AMP-PD (https://amp-pd.org/). All original code has been deposited at GitHub (https://github.com/Ibanez-Lab/) and is publicly available as of the date of publication.

## Acknowledgments

We thank all the participants and their families along with the institutions and all the staff who provided plasma tissue, without whom this study would not have been possible. This work was supported by access to equipment made possible by the Hope Center for Neurological Disorders, the Neurogenomics and Informatics Center (NGI: https://neurogenomics.wustl.edu/) and the Departments of Neurology and Psychiatry at Washington University School of Medicine.

Data used in the preparation of this article were obtained from the Accelerating Medicine Partnership® (AMP®) Parkinson’s Disease (AMP PD) Knowledge Platform. For up-to-date information on the study, visit https://www.amp-pd.org. The AMP® PD program is a public-private partnership managed by the Foundation for the National Institutes of Health and funded by the National Institute of Neurological Disorders and Stroke (NINDS) in partnership with the Aligning Science Across Parkinson’s (ASAP) initiative; Celgene Corporation, a subsidiary of Bristol-Myers Squibb Company; GlaxoSmithKline plc (GSK); The Michael J. Fox Foundation for Parkinson’s Research ; Pfizer Inc.; AbbVie Inc.; Sanofi US Services Inc.; and Verily Life Sciences. ACCELERATING MEDICINES PARTNERSHIP and AMP are registered service marks of the U.S. Department of Health and Human Services.

Clinical data and biosamples used in preparation of this article were obtained from the Michael J. Fox Foundation for Parkinson’s Research (MJFF) Parkinson’s Progression Markers Initiative (PPMI). PPMI is sponsored by The Michael J. Fox Foundation for Parkinson’s Research and supported by a consortium of scientific partners: 4D Pharma, Abbvie, AcureX, Allergan, Amathus Therapeutics, Aligning Science Across Parkinson’s, AskBio, Avid Radiopharmaceuticals, BIAL, BioArctic, Biogen, Biohaven, BioLegend, BlueRock Therapeutics, Bristol-Myers Squibb, Calico Labs, Capsida Biotherapeutics, Celgene, Cerevel Therapeutics, Coave Therapeutics, DaCapo Brainscience, Denali, Edmond J. Safra Foundation, Eli Lilly, Gain Therapeutics, GE HealthCare, Genentech, GSK, Golub Capital, Handl Therapeutics, Insitro, Jazz Pharmaceuticals, Johnson & Johnson Innovative Medicine, Lundbeck, Merck, Meso Scale Discovery, Mission Therapeutics, Neurocrine Biosciences, Neuron23, Neuropore, Pfizer, Piramal, Prevail Therapeutics, Roche, Sanofi, Servier, Sun Pharma Advanced Research Company, Takeda, Teva, UCB, Vanqua Bio, Verily, Voyager v. 25MAR2024 Therapeutics, the Weston Family Foundation and Yumanity Therapeutics. The PPMI investigators have not participated in reviewing the data analysis or content of the manuscript. For up-to-date information on the study, visit www.ppmi-info.org.

## Author contributions

LI, RK and AB conceived and wrote this article. LI conceptualized and designed the research plan. LI, RK, AB, and CC designed the analysis plan. RK, AB, DW, ZY, WL and JT processed all the data and performed the analyses. RK, AB, DW, ZY, WL, JT, CC, and LI discussed the project, revised the manuscript, and provided critical feedback.

## References

1. Vidovic M, Rikalovic MG: Alpha-Synuclein Aggregation Pathway in Parkinson’s Disease: Current Status and Novel Therapeutic Approaches. Cells 2022, 11.

2. Beric A, Sun Y, Sanchez S, Martin C, Powell T, Kumar R, Pardo JA, Darekar G, Sanford J, Dikec D, et al: Circulating blood circular RNA in Parkinson’s Disease; from involvement in pathology to diagnostic tools in at-risk individuals. NPJ Parkinsons Dis 2024, 10:222.

3. Kaiser S, Zhang L, Mollenhauer B, Jacob J, Longerich S, Del-Aguila J, Marcus J, Raghavan N, Stone D, Fagboyegun O, et al: A proteogenomic view of Parkinson’s disease causality and heterogeneity. NPJ Parkinsons Dis 2023, 9:24.

4. Irmady K, Hale CR, Qadri R, Fak J, Simelane S, Carroll T, Przedborski S, Darnell RB: Blood transcriptomic signatures associated with molecular changes in the brain and clinical outcomes in Parkinson’s disease. Nat Commun 2023, 14:3956.

5. Poewe W, Seppi K, Tanner CM, Halliday GM, Brundin P, Volkmann J, Schrag AE, Lang AE: Parkinson disease. Nat Rev Dis Primers 2017, 3:17013.

6. Tolosa E, Garrido A, Scholz SW, Poewe W: Challenges in the diagnosis of Parkinson’s disease. Lancet Neurol 2021, 20:385–397.

7. You J, Wang L, Wang Y, Kang J, Yu J, Cheng W, Feng J: Prediction of Future Parkinson Disease Using Plasma Proteins Combined With Clinical-Demographic Measures. Neurology 2024, 103:e209531.

8. Rutledge J, Lehallier B, Zarifkar P, Losada PM, Shahid-Besanti M, Western D, Gorijala P, Ryman S, Yutsis M, Deutsch GK, et al: Comprehensive proteomics of CSF, plasma, and urine identify DDC and other biomarkers of early Parkinson’s disease. Acta Neuropathol 2024, 147:52.

9. Winchester L, Barber I, Lawton M, Ash J, Liu B, Evetts S, Hopkins-Jones L, Lewis S, Bresner C, Malpartida AB, et al: Identification of a possible proteomic biomarker in Parkinson’s disease: discovery and replication in blood, brain and cerebrospinal fluid. Brain Commun 2023, 5:fcac343.

10. Tonges L, Buhmann C, Klebe S, Klucken J, Kwon EH, Muller T, Pedrosa DJ, Schroter N, Riederer P, Lingor P: Blood-based biomarker in Parkinson’s disease: potential for future applications in clinical research and practice. J Neural Transm (Vienna) 2022, 129:1201–1217.

11. Ma ZL, Wang ZL, Zhang FY, Liu HX, Mao LH, Yuan L: Biomarkers of Parkinson’s Disease: From Basic Research to Clinical Practice. Aging Dis 2024, 15:1813–1830.

12. Siderowf A, Concha-Marambio L, Lafontant DE, Farris CM, Ma Y, Urenia PA, Nguyen H, Alcalay RN, Chahine LM, Foroud T, et al: Assessment of heterogeneity among participants in the Parkinson’s Progression Markers Initiative cohort using alpha-synuclein seed amplification: a cross-sectional study. Lancet Neurol 2023, 22:407–417.

13. Berg D, Klein C: alpha-synuclein seed amplification and its uses in Parkinson’s disease. Lancet Neurol 2023, 22:369–371.

14. Eldjarn GH, Ferkingstad E, Lund SH, Helgason H, Magnusson OT, Gunnarsdottir K, Olafsdottir TA, Halldorsson BV, Olason PI, Zink F, et al: Large-scale plasma proteomics comparisons through genetics and disease associations. Nature 2023, 622:348–358.

15. Dammer EB, Ping L, Duong DM, Modeste ES, Seyfried NT, Lah JJ, Levey AI, Johnson ECB: Multi-platform proteomic analysis of Alzheimer’s disease cerebrospinal fluid and plasma reveals network biomarkers associated with proteostasis and the matrisome. Alzheimers Res Ther 2022, 14:174.

16. Katz DH, Robbins JM, Deng S, Tahir UA, Bick AG, Pampana A, Yu Z, Ngo D, Benson MD, Chen ZZ, et al: Proteomic profiling platforms head to head: Leveraging genetics and clinical traits to compare aptamer- and antibody-based methods. Sci Adv 2022, 8:eabm5164.

17. Raffield LM, Dang H, Pratte KA, Jacobson S, Gillenwater LA, Ampleford E, Barjaktarevic I, Basta P, Clish CB, Comellas AP, et al: Comparison of Proteomic Assessment Methods in Multiple Cohort Studies. Proteomics 2020, 20:e1900278.

18. Pietzner M, Wheeler E, Carrasco-Zanini J, Kerrison ND, Oerton E, Koprulu M, Luan J, Hingorani AD, Williams SA, Wareham NJ, Langenberg C: Synergistic insights into human health from aptamer- and antibody-based proteomic profiling. Nat Commun 2021, 12:6822.

19. Hallqvist J, Bartl M, Dakna M, Schade S, Garagnani P, Bacalini MG, Pirazzini C, Bhatia K, Schreglmann S, Xylaki M, et al: Plasma proteomics identify biomarkers predicting Parkinson’s disease up to 7 years before symptom onset. Nat Commun 2024, 15:4759.

20. Jang Y, Pletnikova O, Troncoso JC, Pantelyat AY, Dawson TM, Rosenthal LS, Na CH: Mass Spectrometry-Based Proteomics Analysis of Human Substantia Nigra From Parkinson’s Disease Patients Identifies Multiple Pathways Potentially Involved in the Disease. Mol Cell Proteomics 2023, 22:100452.

21. Shao Y, Li T, Liu Z, Wang X, Xu X, Li S, Xu G, Le W: Comprehensive metabolic profiling of Parkinson’s disease by liquid chromatography-mass spectrometry. Mol Neurodegener 2021, 16:4.

22. Blumenreich S, Nehushtan T, Kupervaser M, Shalit T, Gabashvili A, Joseph T, Milenkovic I, Hardy J, Futerman AH: Large-scale proteomics analysis of five brain regions from Parkinson’s disease patients with a GBA1 mutation. NPJ Parkinsons Dis 2024, 10:33.

23. Karayel O, Virreira Winter S, Padmanabhan S, Kuras YI, Vu DT, Tuncali I, Merchant K, Wills AM, Scherzer CR, Mann M: Proteome profiling of cerebrospinal fluid reveals biomarker candidates for Parkinson’s disease. Cell Rep Med 2022, 3:100661.

24. Virreira Winter S, Karayel O, Strauss MT, Padmanabhan S, Surface M, Merchant K, Alcalay RN, Mann M: Urinary proteome profiling for stratifying patients with familial Parkinson’s disease. EMBO Mol Med 2021, 13:e13257.

25. Rotunno MS, Lane M, Zhang W, Wolf P, Oliva P, Viel C, Wills AM, Alcalay RN, Scherzer CR, Shihabuddin LS, et al: Cerebrospinal fluid proteomics implicates the granin family in Parkinson’s disease. Sci Rep 2020, 10:2479.

26. Dong W, Qiu C, Gong D, Jiang X, Liu W, Liu W, Zhang L, Zhang W: Proteomics and bioinformatics approaches for the identification of plasma biomarkers to detect Parkinson’s disease. Exp Ther Med 2019, 18:2833–2842.

27. Posavi M, Diaz-Ortiz M, Liu B, Swanson CR, Skrinak RT, Hernandez-Con P, Amado DA, Fullard M, Rick J, Siderowf A, et al: Characterization of Parkinson’s disease using blood-based biomarkers: A multicohort proteomic analysis. PLoS Med 2019, 16:e1002931.

28. Abdi IY, Bartl M, Dakna M, Abdesselem H, Majbour N, Trenkwalder C, El-Agnaf O, Mollenhauer B: Cross-sectional proteomic expression in Parkinson’s disease-related proteins in drug-naive patients vs healthy controls with longitudinal clinical follow-up. Neurobiol Dis 2023, 177:105997.

29. Ibanez L, Bahena JA, Yang C, Dube U, Farias FHG, Budde JP, Bergmann K, Brenner-Webster C, Morris JC, Perrin RJ, et al: Functional genomic analyses uncover APOE-mediated regulation of brain and cerebrospinal fluid beta-amyloid levels in Parkinson disease. Acta Neuropathol Commun 2020, 8:196.

30. Shi X, Wei T, Hu Y, Wang M, Tang Y: The associations between plasma soluble Trem1 and neurological diseases: a Mendelian randomization study. J Neuroinflammation 2022, 19:218.

31. Paslawski W, Khosousi S, Hertz E, Markaki I, Boxer A, Svenningsson P: Large-scale proximity extension assay reveals CSF midkine and DOPA decarboxylase as supportive diagnostic biomarkers for Parkinson’s disease. Transl Neurodegener 2023, 12:42.

32. Bartl M, Dakna M, Schade S, Otte B, Wicke T, Lang E, Starke M, Ebentheuer J, Weber S, Toischer K, et al: Blood Markers of Inflammation, Neurodegeneration, and Cardiovascular Risk in Early Parkinson’s Disease. Mov Disord 2023, 38:68–81.

33. Santaella A, Kuiperij HB, van Rumund A, Esselink RAJ, van Gool AJ, Bloem BR, Verbeek MM: Inflammation biomarker discovery in Parkinson’s disease and atypical parkinsonisms. BMC Neurol 2020, 20:26.

34. Bolsewig K, Willemse EAJ, Sanchez-Juan P, Rabano A, Martinez M, Doecke JD, Bellomo G, Vermunt L, Alcolea D, Halbgebauer S, et al: Increased plasma DOPA decarboxylase levels in Lewy body disorders are driven by dopaminergic treatment. Nat Commun 2025, 16:1139.

35. Pereira JB, Kumar A, Hall S, Palmqvist S, Stomrud E, Bali D, Parchi P, Mattsson-Carlgren N, Janelidze S, Hansson O: DOPA decarboxylase is an emerging biomarker for Parkinsonian disorders including preclinical Lewy body disease. Nat Aging 2023, 3:1201–1209.

36. Gan YH, Ma LZ, Zhang Y, You J, Guo Y, He Y, Wang LB, He XY, Li YZ, Dong Q, et al: Large-scale proteomic analyses of incident Parkinson’s disease reveal new pathophysiological insights and potential biomarkers. Nat Aging 2025.

37. Tsukita K, Sakamaki-Tsukita H, Kaiser S, Zhang L, Messa M, Serrano-Fernandez P, Takahashi R: High-Throughput CSF Proteomics and Machine Learning to Identify Proteomic Signatures for Parkinson Disease Development and Progression. Neurology 2023, 101:e1434–e1447.

38. Funayama M, Nishioka K, Li Y, Hattori N: Molecular genetics of Parkinson’s disease: Contributions and global trends. J Hum Genet 2023, 68:125–130.

39. Pitz V, Makarious MB, Bandres-Ciga S, Iwaki H, andMe Research T, Singleton AB, Nalls M, Heilbron K, Blauwendraat C: Analysis of rare Parkinson’s disease variants in millions of people. NPJ Parkinsons Dis 2024, 10:11.

40. Li Y, Kang W, Zhang L, Zhou L, Niu M, Liu J: Hyposmia Is Associated with RBD for PD Patients with Variants of SNCA. Front Aging Neurosci 2017, 9:303.

41. Kawabata K, Bagarinao E, Seppi K, Poewe W: Longitudinal brain changes in Parkinson’s disease with severe olfactory deficit. Parkinsonism Relat Disord 2024, 122:106072.

42. Yu G, Wang LG, Han Y, He QY: clusterProfiler: an R package for comparing biological themes among gene clusters. OMICS 2012, 16:284–287.

43. Bates D, Mächler, M., Bolker, B., & Walker, S.: Fitting Linear Mixed-Effects Models Using lme4. Journal of Statistical Software 2015, 67:1–48.

44. Chang CC, Chow CC, Tellier LC, Vattikuti S, Purcell SM, Lee JJ: Second-generation PLINK: rising to the challenge of larger and richer datasets. Gigascience 2015, 4:7.

45. Appleton E, Khosousi S, Ta M, Nalls M, Singleton AB, Sturchio A, Markaki I, Paslawski W, Iwaki H, Svenningsson P: DOPA-decarboxylase is elevated in CSF, but not plasma, in prodromal and de novo Parkinson’s disease. Transl Neurodegener 2024, 13:31.

46. Wang B, Pozarickij A, Mazidi M, Wright N, Yao P, Said S, Iona A, Kartsonaki C, Fry H, Lin K, et al: Comparative studies of 2168 plasma proteins measured by two affinity-based platforms in 4000 Chinese adults. Nat Commun 2025, 16:1869.

47. Nahnsen S, Bielow C, Reinert K, Kohlbacher O: Tools for label-free peptide quantification. Mol Cell Proteomics 2013, 12:549–556.

48. Birhanu AG: Mass spectrometry-based proteomics as an emerging tool in clinical laboratories. Clin Proteomics 2023, 20:32.

