## Supplementary Figures for "Orthogonal validation of PD Biomarkers: Multi-platform proteomics profiling of CSF, Plasma, and Urine confirms DDC as a consistent candidate"

*Equal authors

**^#^Correspondence:**

Laura Ibanez, PhD

Washington University in Saint Louis School of Medicine

4444 Forest Park Ave

Campus Box 8134

Saint Louis, MO 63110

**Supplementary Figure 1.** Upset plots showing the number of overlapping (A) samples across different platforms and tissues, (B) individuals across different platforms and tissues.

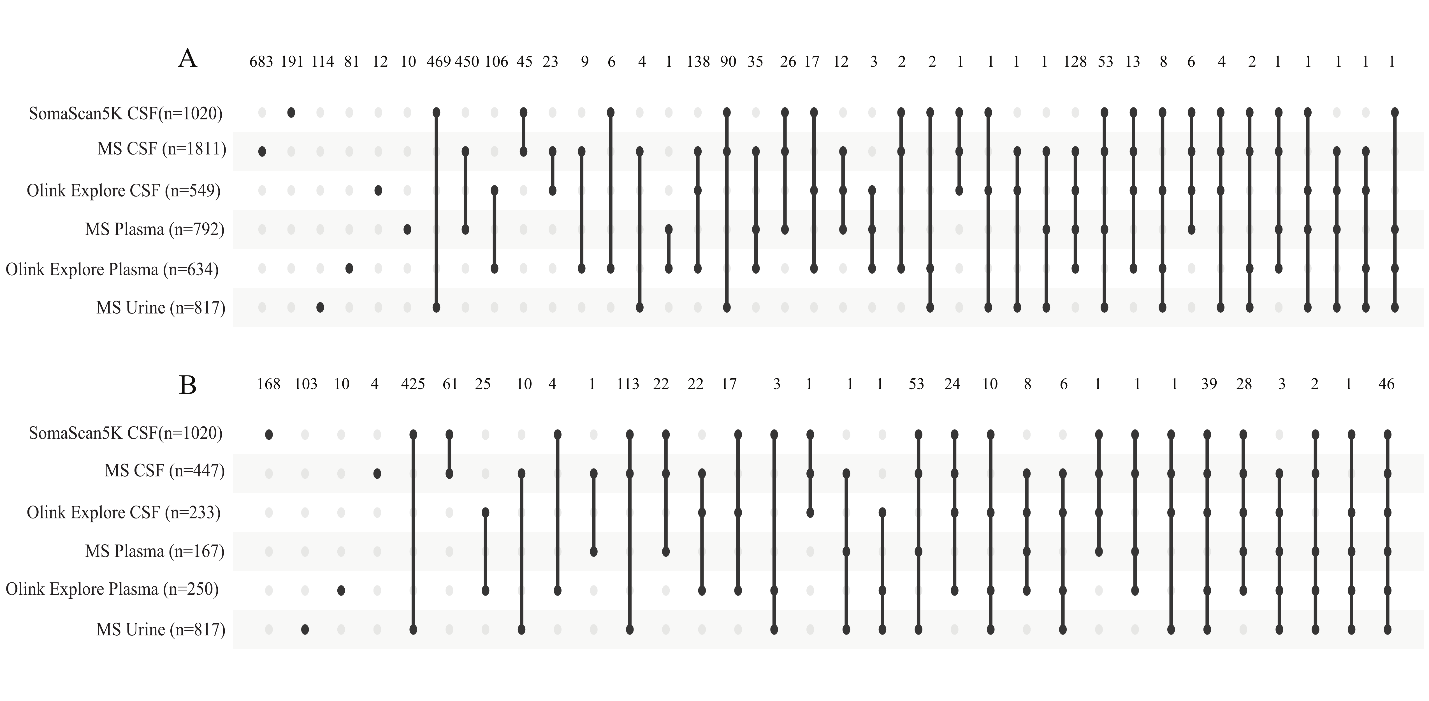

**Supplementary Figure 2.** Data distribution before and after z-score scaling. Panels A and B show CSF from SomaScan5K, panels C and D shows CSF from MS, panels E and F show plasma from MS, and panels G and H show urine from MS before (left) and after (right) z-score scaling.

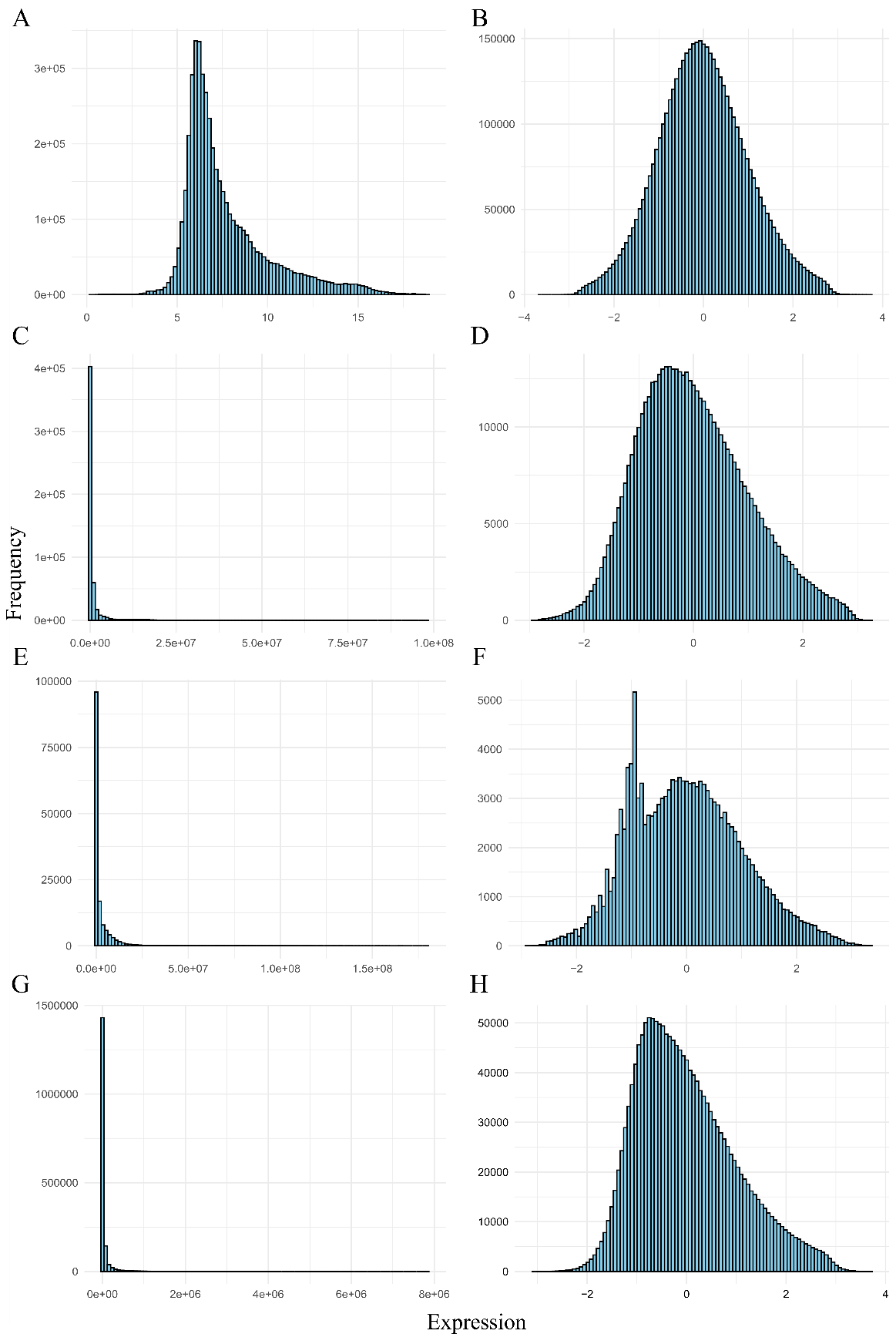

**Supplementary Figure 3.** Quality control summary. The scatter plots represent the principal components of all the measurements that survived quality control for CSF samples quantified with (A) SomaScan5K, (B) MS, or (C) Olink Explore, plasma samples quantified with (D) MS or (E) Olink Explore and (F) urine samples quantified with MS.

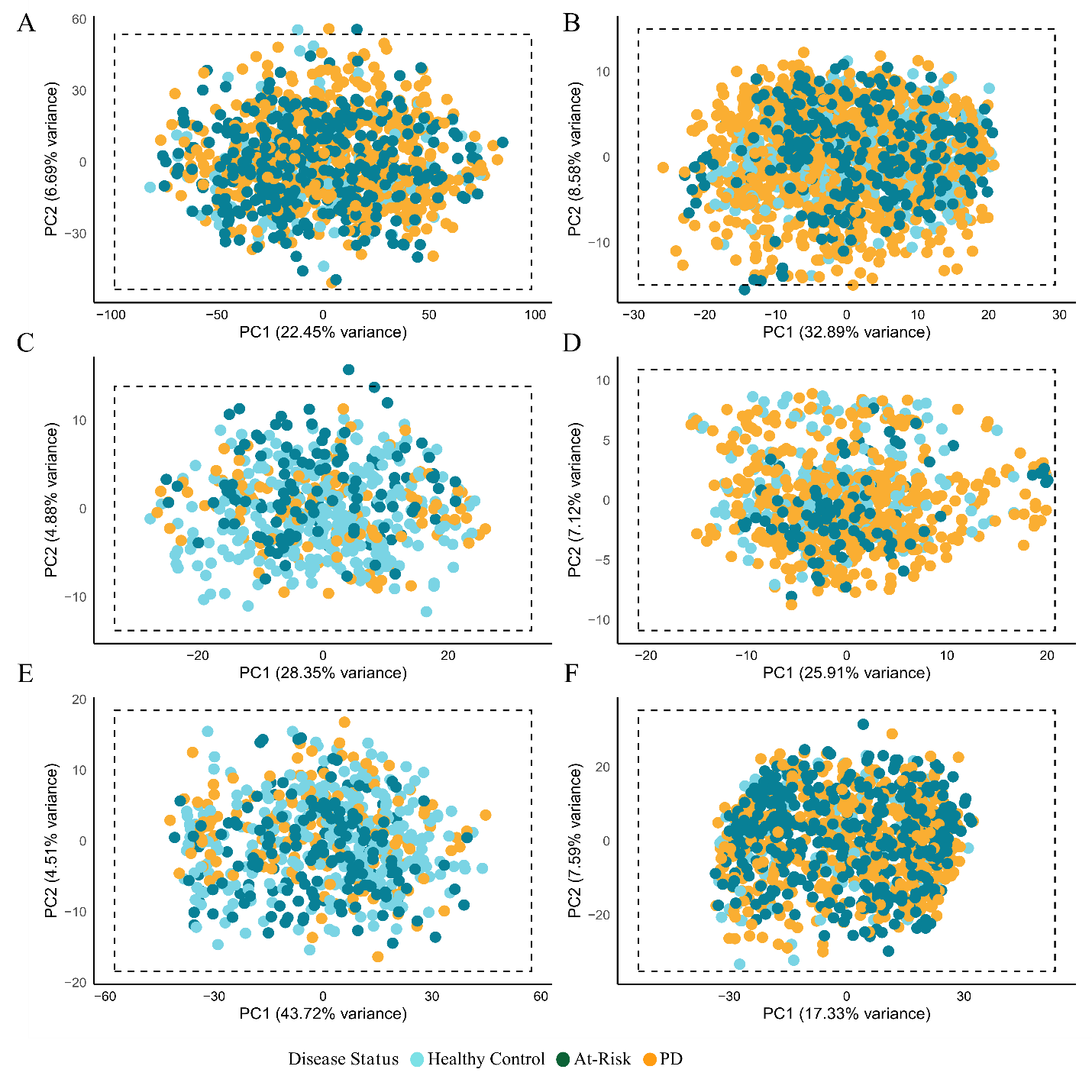

**Supplementary Figure 4.** Scatter plots showing correlations of beta values (effect size) from baseline measurements between **(A)** 71 proteins quantified in CSF by SomaScan5K and MS; **(B)** 373 proteins quantified in CSF using SomaScan5K and Olink Explore **(C)** 71 proteins quantified in CSF by MS and Olink Explore **(D)** 20 proteins quantified in plasma using MS and Olink Explore.

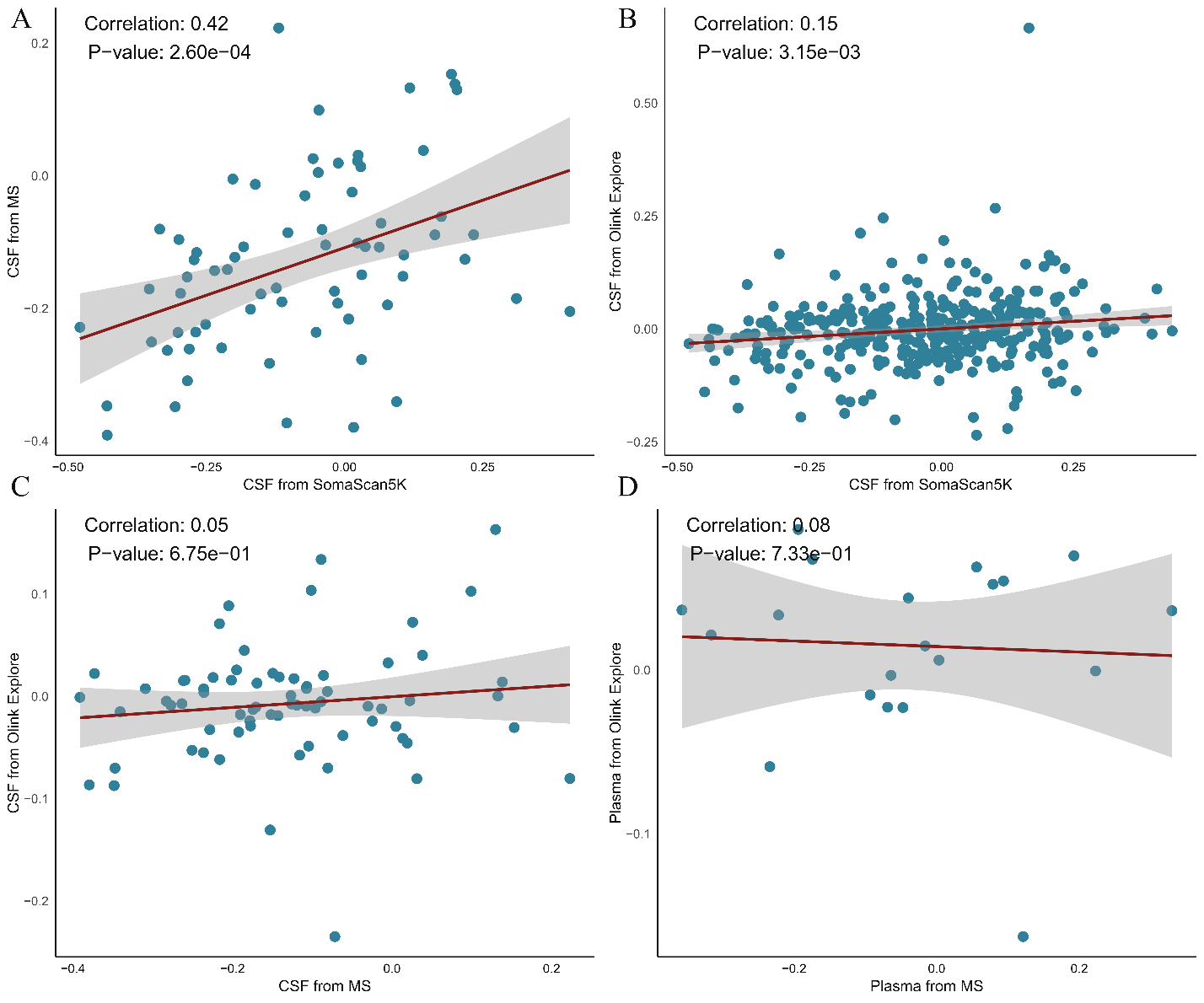

**Supplementary Figure 5.** Violin plot showing the expression of DDC in healthy control, at-risk, and PD participants in (A) CSF from Olink Explore, (B) urine from MS. X and Y-axis shows disease status and DDC expression respectively.

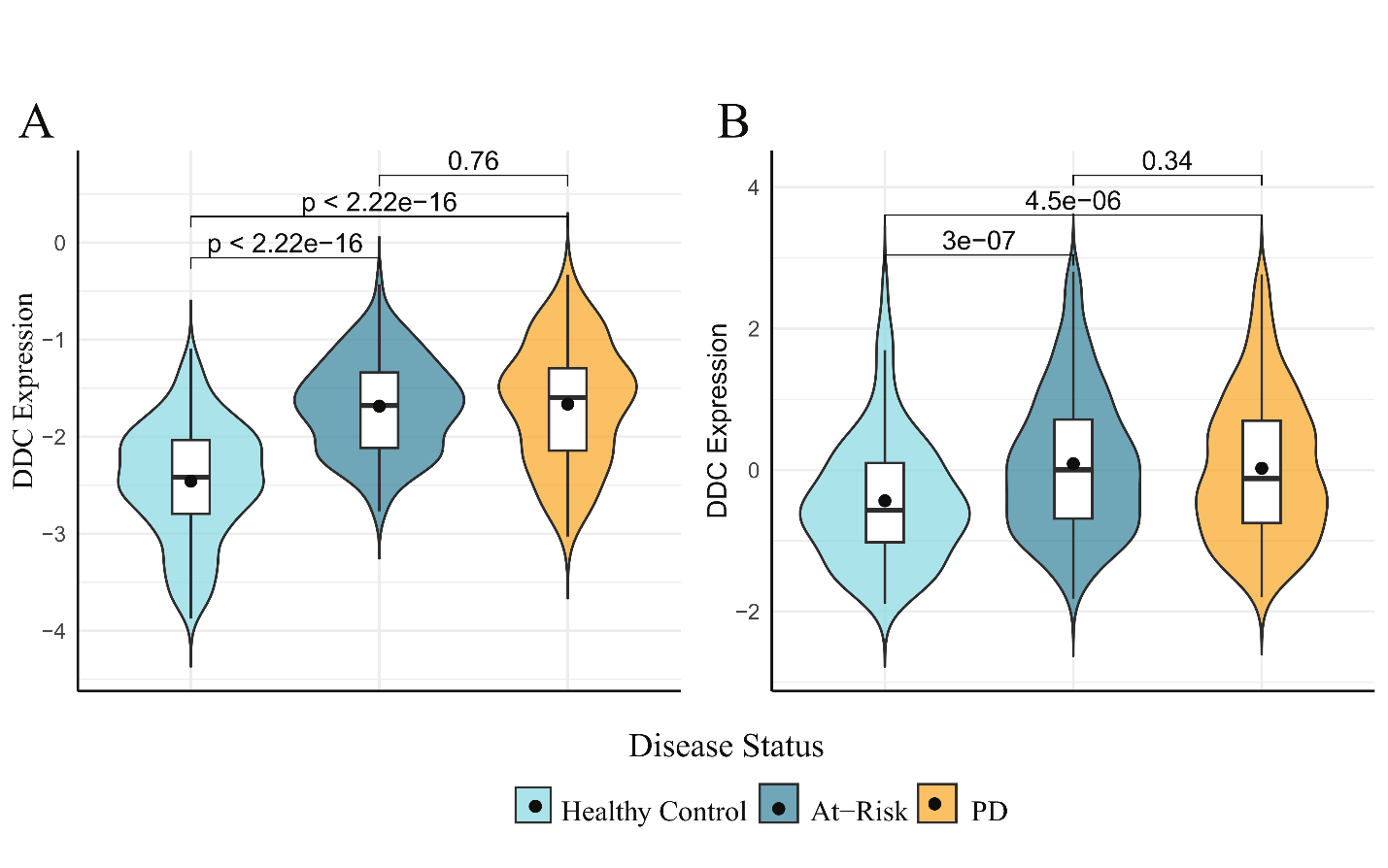

**Supplementary Figure 6.** DDC expression over time in CSF from Olink Explore.

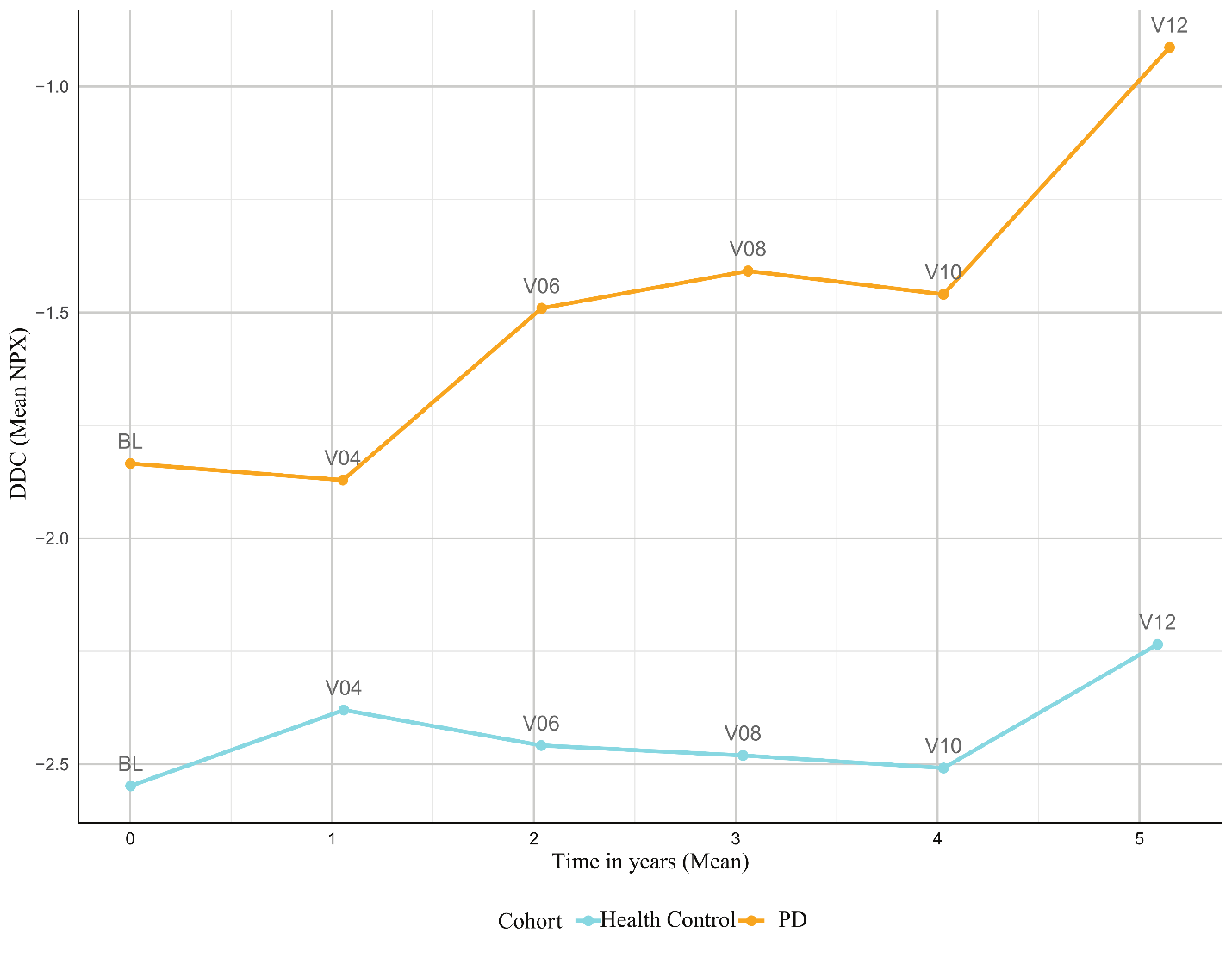

**Supplementary Figure 7.** Volcano plots showing the results of baseline differential accumulation analyses in CSF from (A) SomaScan5K, (B) MS, (C) Olink Explore; in plasma from (D) MS, and (E) Olink Explore; and in urine from (F) MS. X and Y-axis represents beta and -log10(p-value) respectively. Yellow color represents multiple test corrected proteins while gray color shows non-significant proteins.

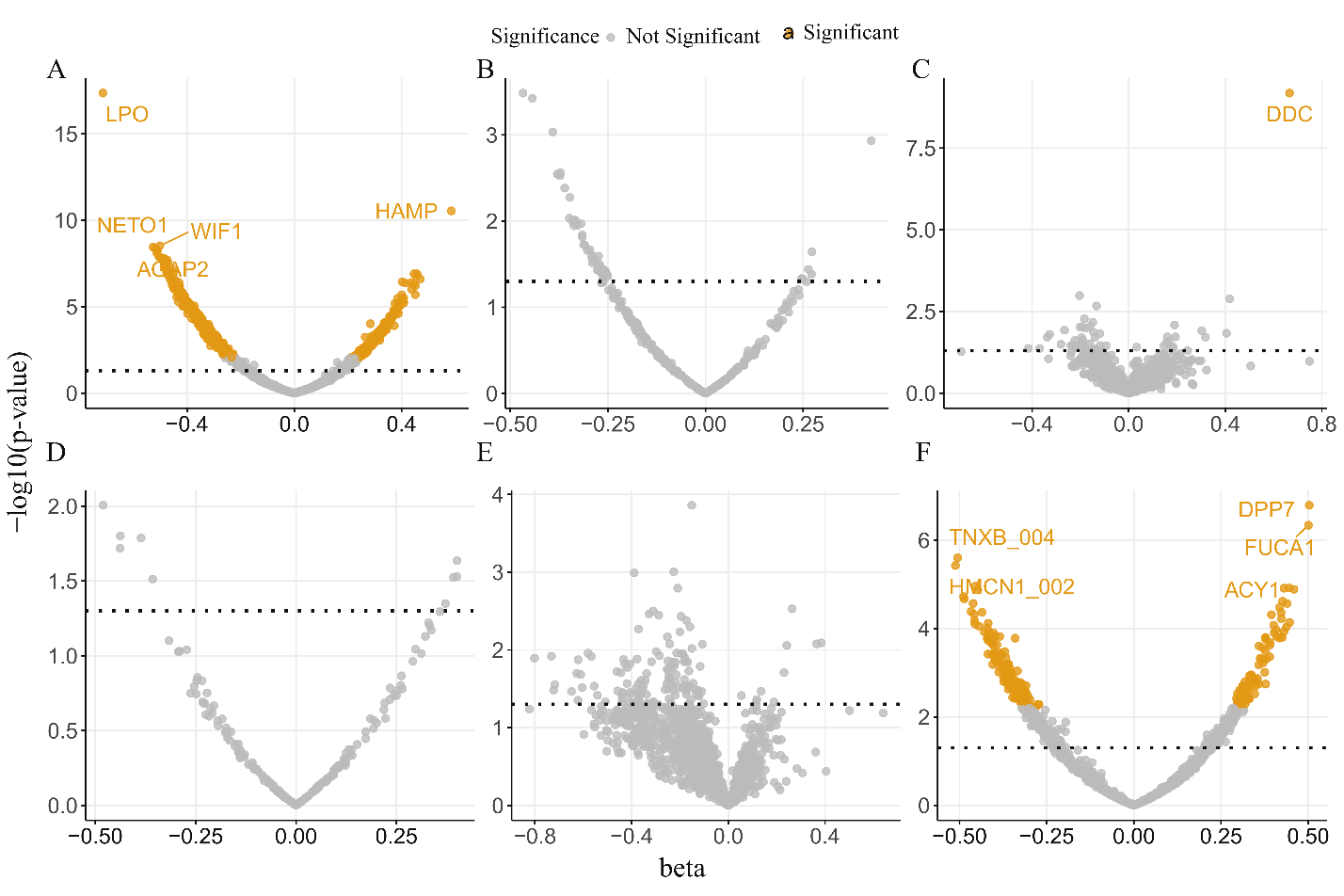
